# In-person schooling and COVID-19 transmission in Canada’s three largest cities

**DOI:** 10.1101/2021.03.21.21254064

**Authors:** Simona Bignami-van Assche, Yacine Boujija, David Fisman, John Sandberg

**Affiliations:** Département de démographie, Université de Montréal; Epidemiology Division, University of Toronto; School of Public Health, George Washington University

## Abstract

In North America and Europe, the Fall 2020 school term has coincided with the beginning of the second wave of the novel coronavirus (COVID-19) pandemic, sparking a heated debate about the role of in-person schooling for community transmission of severe acute respiratory syndrome coronavirus 2 (SARS-CoV-2). This issue has immediate policy relevance for deciding how to operate schools safely as the pandemic unfolds, new variants of SARS-CoV-2 are circulating, and immunization coverage remains low. We contribute to this debate by presenting data on trends in COVID-19 weekly incidence among school-aged children 0-19 years old vis-à-vis other age groups during Fall 2020 in Canada’s three largest cities: Montréal, Toronto and Calgary. We interpret these trends in light of the different back-to-school policies and other public health measures implemented in the three cities over the observation period.

**KEY POINTS:**

- School closures are an effective measure to reduce the overall incidence of the novel coronavirus (COVID-19). Nonetheless, there is a general consensus that the decision to close schools to control the spread of COVID-19 should be used as last resort because of the negative impact on children’s development and mental health, and since they are less likely to have severe COVID-19 outcomes than adults.
- Existing evidence highlights the importance of adopting appropriate mitigation strategies for limiting COVID-19 community spread when returning to in-person schooling. To understand the association between in-person schooling and COVID-19 transmission given different mitigation strategies, especially universal masking and distance learning, we compare how the second wave of COVID-19 has affected school-aged children age 0-19 years old vis-à-vis other age groups in Montréal, Toronto and Calgary during Fall 2020.
- The case of Montréal attests to the negative consequences of not implementing recommended migration strategies when reopening schools, even when public health measures such as gatherings restrictions are in place to maintain low levels of community transmission. On the contrary, school measures adopted in Toronto (optional distance learning and masking mandates), have limited the role of COVID-19 transmission among school-aged children for overall community transmission. In Calgary, this effect has been smaller, likely because public health measures to limit COVID-19 community spread were not introduced until early December 2020.
- Our findings have immediate policy relevance for deciding how to operate schools safely as the pandemic unfolds, new variants of SARS-CoV-2 are circulating, and immunization coverage remains low.

---

After prolonged, worldwide school closures during the first wave of the novel coronavirus (COVID-19) pandemic in spring 2020, the Fall school term has marked the return to in-person instruction for millions of children in Europe and North America. However, this period has also coincided with the second pandemic wave in these regions, sparking a heated debate about the role of in-person schooling for community transmission of severe acute respiratory syndrome coronavirus 2 (SARS-CoV-2), which causes COVID-19.

To limit COVID-19 transmission in school settings, existing evidence highlights the importance of maintaining low levels of transmission in the surrounding communities and adopting recommended mitigation strategies such as universal face mask use and hybrid attendance models to limit crowding in classrooms (Honein, Barrios and Brooks, 2021). Nonetheless, this evidence refers primarily to settings with low levels of in-person school attendance and varying levels of community transmission.

In Canada, levels of community transmission were low when schools opened in Fall 2020. On the contrary, back-to-school policies were quite heterogenous, notably with respect to mask mandates and mode of instruction (fully in person / with optional distance learning). Comparing how the second wave of COVID-19 has affected school-aged children and communities in Montréal, Toronto and Calgary during Fall 2020 thus offers an opportunity to understand the association between in-person school attendance and COVID-19 transmission given different mitigation strategies and homogenous levels of pre-existing community transmission.

In this paper, we present data on trends in COVID-19 weekly incidence among school-aged children 0-19 years old vis-à-vis other age groups during Fall 2020 in Canada’s three largest cities: Montréal, Toronto and Calgary. We interpret these trends in light of the different back-to-school policies and other public health measures implemented in the three cities over the observation period. Our findings have immediate policy relevance for deciding how to operate schools safely as the pandemic unfolds, new variants of SARS-CoV-2 are circulating, and immunization coverage remains low.

## Background

School closures are an effective measure in reducing the reproduction number of COVID-19 (Brauner et al., 2020; Li et al., 2020) and the overall incidence of SARS-CoV-2 infections (ECDC, 2020). Nonetheless, prolonged school closures have serious negative implications for children’s health and development (Kuhfeld et al. 2020; Hertz and Barrios, 2020; ECDC, 2020; Chanchlani et al., 2020). For this latter reason, in Fall 2020 in-person schooling and avoiding large-scale school closures have been a priority in Canada as elsewhere.

Return to in-person schooling has sparked a heated debate about how schools can operate safely as the pandemic unfolds. As embedded in the World Health Organization’s recommendations, two main conditions need to be satisfied to do so: community transmission of COVID-19 should be maintained at low levels through appropriate policy measures (such as restrictions on indoor gatherings); and recommended mitigation strategies in schools should be in place (WHO, 2020a; 2020b). These measures include cohorting (keeping students and staff in small groups or “bubbles” that do not mix); the universal use of masks; limiting physical presence in classroom in order to reduce crowding; and ensuring adequate and appropriate ventilation.

Large-scale, population-based studies on the role of these conditions for the spread of SARS-CoV-2 between school settings and surrounding communities come primarily from the United States (Honein, Barrios and Brooks, 2020). Notably, given almost universal face mask use and low levels of exclusively in-person schooling (less than 20 percent of school districts), a recent national assessment found that increases in COVID-19 incidence and percentage of positive test results among adults were not preceded by increases in the school-aged population 0-24 years old (US Department of Health and Human Services / Center for Disease Control, 2020).^1^ In other terms, levels of COVID-19 transmission in school settings were not a contributing factor of community transmission, but rather reflected it given that recommended mitigation strategies were widely adopted. Similar findings applied to settings with varying levels of pre-existing community transmission (Goldhaber et al., 2020).

Since only a small proportion of US primary and secondary students attended school exclusively in person during Fall 2020, it is currently unknown how the association between in-person school attendance and COVID-19 transmission varies when different mitigation strategies (notably universal face mask use and distance/hybrid learning) are in place. We shed light on this issue by presenting data on trends in COVID-19 weekly incidence among school-aged children 0-19 years old vis-à-vis other age groups during Fall 2020 in Canada’s three largest cities: Montréal, Toronto and Calgary.^2^

Although levels of COVID-19 community transmission were low at the beginning of the school year in the three cities considered, back-to-school plans were quite different. Cohorting was the main common element, whereas the mode of instruction and mask mandates differed. Provincial plans (Toronto) or school boards’ plans (Calgary) allowed for distance learning upon parents’ request. Montréal followed the only provincial plan in Canada requiring all children to attend school in person with limited exemptions granted exclusively in case of severe health risks.^3^ As for mask mandates, in Ontario masks were required in classrooms and common areas in both elementary and secondary schools in the largest public health units in the province, including Toronto, Ottawa, Peel and York, and were encouraged in kindergarten in these health units as well. In Montréal, masks have been mandatory only in common areas in elementary schools for the Fall semester, and they became mandatory in classrooms as well in secondary schools in early October. In Calgary, masks were mandatory from kindergarden to Grade 12 since the beginning of the Fall semester but could be removed by students when seated in classroom with their cohorts and physical distance was ensured.

## Data and Methods

Official counts of COVID-19 cases (defined as positive real-time reverse transcription– polymerase chain) by age were obtained from provincial reporting jurisdictions. For Montréal, they were extracted from aggregate counts of COVID-19 cases released in weekly reports by the Direction régionale de santé publique^4^. For Toronto and Calgary, individual-level case report data were available from, respectively, Ontario Health^5^ and Alberta Health^6^.

Following a standard methodology (US Department of Health and Human Services / Center for Disease Control, 2020), COVID-19 case data were analyzed to describe trends in weekly incidence of the disease among children, adolescents and young adults (aged 0-19 years) between August 18, 2020 and January 12, 2021. Due to data availability, children and young people could be stratified only in two age groups (0-9 and 10-19 years old), which do not align perfectly with educational groupings (kindergarden, primary and secondary). COVID-19 weekly incidence is expressed as cases per 100,000. Population estimates used in calculating incidence were obtained from Statistics Canada (Statistics Canada, 2020).

### How has COVID-19 incidence in 0-19 years old evolved in Montréal, Toronto and Calgary?

At the end of August 2020, weekly reported incidence of COVID-19 was under 30 per 100,000 in Montréal, Toronto and Calgary (see Table 1). Public health measures to maintain low levels of community transmission (notably restrictions on mass and social gatherings) were implemented in Montréal and Toronto in early October 2020. As a result, the second wave of COVID-19 hit first and progressed faster in Calgary than in the other two cities, where it picked up speed only in late October. However, Alberta implemented strict public health measures to reduce community transmission in early December 2020 so that, for the week of December 29 to January 4, COVID-19 weekly incidence was 356.9 cases per 100,000 in Montréal, 165.9 cases per 100,000 in Toronto, and 153.5 cases per 100,000 in Calgary.

**Table 1:**
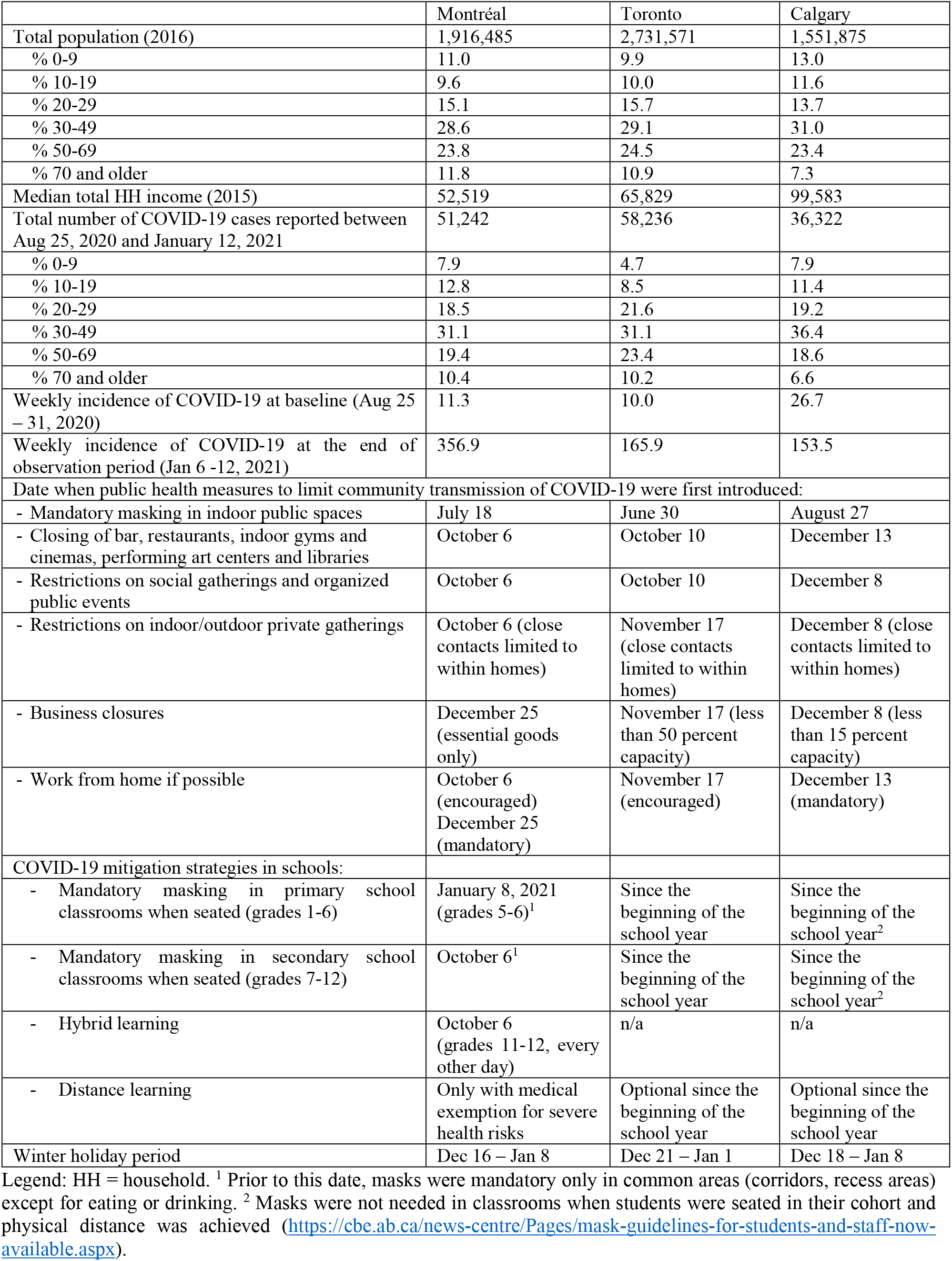
Demographics and pandemic indicators for Montréal, Toronto and Calgary

In all three cities, at the end of December 2020 the largest proportion of confirmed SARS-CoV-2 infections was found among 30-59 years old, followed by 0-19 years old. The proportion of confirmed infections under age 20 was higher in Québec than in Alberta, even though this subgroup represents a larger share of the population in the former than the latter case (see Table 1). Indeed, the distribution of COVID-19 cases by age in each city at the end of the 2020 Fall term results from the evolution of weekly incidence in different age groups (Figure 1). In Toronto and Calgary, trends in children and adolescents aged 0-19 years paralleled those among adults. On the contrary, in Montréal increases in incidence among adults 30-49 years old were preceded by increases among school-aged children, especially those aged 10-19 years. These increases were little impacted by the mitigation strategies introduced in schools in early October, which included mask mandates in secondary school classrooms and the interruption of sports and extracurricular activities.^7^ Once COVID-19 transmission among 10-19 years old ‘spilled over’ to 30-49 years old, we observe increases in COVID-19 incidence across all other age groups, resulting in an exponential increase of the overall incidence of the disease. In Montréal, transmission of COVID-19 in school-aged children thus does not seem the consequence of community transmission because, in that case, we would expect an increase in incidence for 10-19 years old at the same rate as the incidence in 30-49 years old, like we observe in Toronto and Calgary.

**Figure 1.**
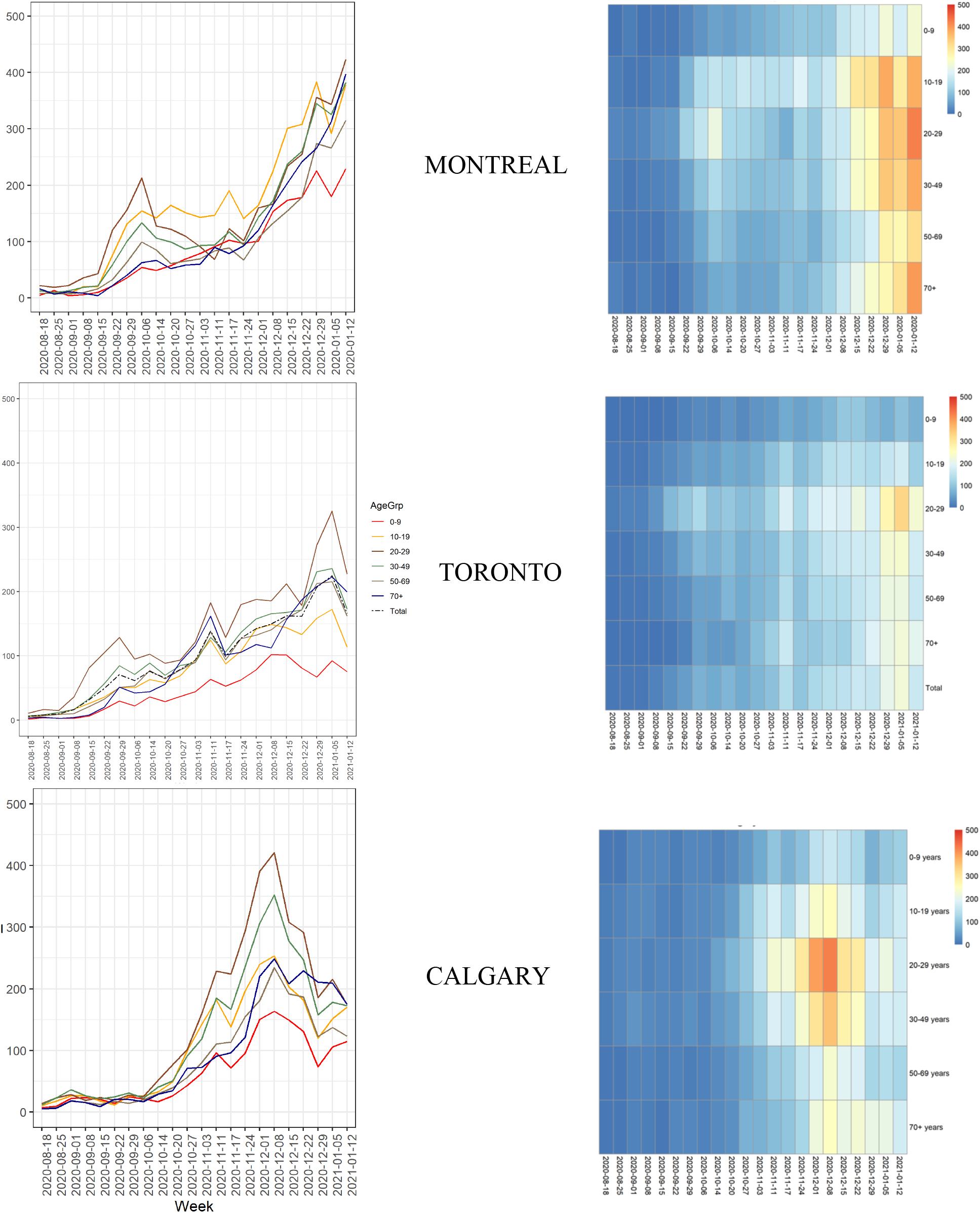
Weekly incidence of COVID-19 in Montréal, Toronto and Calgary by broad age groups: August 18, 2020 – January 12, 2021

### What has been the role of in-person schooling for COVID-19 trends in Montréal, Toronto and Calgary?

The evolution of COVID-19 weekly incidence for 0-19 vs 30 years and older during Fall 2020 in Toronto is consistent with evidence from the US (US Department of Health and Human Services / Center for Disease Control, 2020), where mitigation strategies in schools, notably universal mask mandates and distance/hybrid learning, have been similar. The case of Calgary underscores the importance of optional distance learning for limiting spillover effects from school-aged children to other age groups. Indeed, in spite of the lack of public health measures to limit community transmission of COVID-19 for most of the Fall semester, in Calgary weekly incidence in 0-19 years has mirrored transmission in other age groups, notably 30-49 years old, like in Toronto. Montréal provides an extreme example of the negative consequences on COVID-19 incidence of not implementing recommended migration strategies when reopening schools, even when public health measures such as gatherings restrictions are in place to maintain low levels of community transmission. Our findings for Montréal are in line with data from the UK Office for National Statistics, which shows that the prevalence of infection among children aged 2–16 years rose above the prevalence for all other age groups before the 2020 Christmas break (Gurdasani et al., 2021).

In Montréal, it could be argued that trends in COVID-19 incidence among school-aged children may overstate the role of in-person schooling since they might reflect transmission in this age group occurring outside school walls, for instance during sport activities and other social gatherings. This is unlikely to be the case for a number of reasons, however. First, results from an ongoing study on social contacts indicates that, when they reopened in September 2020, schools became the main place of interaction for children and adolescents age 0-19 (INSPQ, 2021). Second, as indicated earlier, extracurricular activities were suspended, and indoor private gatherings were prohibited after early October 2020. Third, in related work, we have identified that Montreal boroughs with the highest incidence of COVID-19 among 0-19 years old are located in health regions with the highest number of schools reporting cases of COVID-19 and the highest number of cases of COVID-19 confirmed in these schools (Bignami et al., 2020). Lastly, Figure 1 shows that, one week after schools closed for the Christmas break (between December 22 and 29), COVID-19 weekly incidence declined among 0-9 and 10-19 years old, while it was still rising exponentially in all other age groups.

## Discussion and conclusions

Existing studies indicate that schools can operate safely as the pandemic unfolds by adopting appropriate public health measure to maintain low levels of COVID-19 community transmission and recommended mitigation strategies in schools such as universal masking and hybrid learning. The role of these strategies has been studied at varying levels of community transmission, but at low prevalence of exclusively in-person schooling. Our findings thus complement the available evidence regarding risk for transmission associated with in-person schooling in important ways.

Mandatory in-person schooling without universal mask mandates (which remain limited to children in grade 5 and higher) have contributed to increase community transmission in Montréal. This has been in spite of partial lockdown measures in place since early October 2020, which include the suspension of school sports programs and tight restrictions on mass, social and indoor gatherings (including dining in restaurants). In Toronto, where universal mask mandates in school and optional distance learning have been implemented since the beginning of the Fall semester, we observe no meaningful contribution of in-person schooling to COVID-19 community transmission, even though indoor private gatherings were allowed until late November 2020. In Calgary, we also observe no meaningful contribution of in-person schooling to COVID-19 community transmission, but higher weekly incidence among 0-19 years old than in Toronto, likely because public health measures to limit COVID-19 community spread were not introduced until early December 2020. These findings highlight the importance of minimizing community transmission and ensuring that recommended mitigation strategies are in place in school settings to ensure a safe environment for in-person learning especially at this time, while COVID-19 incidence in school-age children and the prevalence of new COVID-19 variants are rising sharply.

Our study has a number of limitations that should be borne in mind. First, this is a comparative case study, which is inherently limited to a degree due to the complexity of the variation in mitigation measures at all levels between them. We believe, however, that the comparisons are strong enough to support the inferences drawn here concerning the importance of school mitigation strategies in limiting COVID-19 transmission. This is especially because COVID-19 incidence is likely underestimated in children, especially at younger ages, in part because milder illness and a higher fraction of asymptomatic infections are associated with decreased testing and case finding (Fisman et al., 2021). The second study limitation is that, since data were not available for Montréal and Calgary, we could not assess trends in test positivity by age. Finally, we could not evaluate trends in COVID-19 incidence among teachers and school staff members, because in Canada cases are not routinely reported by occupations other than health care workers.

## Data Availability

Official counts of COVID-19 cases (defined as positive real-time reverse transcription-polymerase chain) by age were obtained from provincial reporting jurisdictions. For Montreal, they were extracted from aggregate counts of COVID-19 cases released in weekly reports by the Direction regionale de sante publique. For Toronto and Calgary, individual-level case report data were available from, respectively, Ontario Health and Alberta Health.

https://santemontreal.qc.ca/fileadmin/fichiers/Campagnes/coronavirus/situation-montreal/rapports-etat-evaluation-montreal/COVID19-Situation-Montreal-Arrondissements-VillesLiees.pdf

https://www.alberta.ca/covid-19-alberta-data.aspx

## Acknowledgements

We would like to thank Andrew Noymer, Pierre Vachon, and Ari Van Assche for their helpful comments on earlier drafts of this manuscript.

## Footnotes

1 At the higher-education level, in-person instruction at colleges and universities in the United States during Fall 2020 was associated with increased county-level COVID-19 incidence and percentage test positivity. Non-university counties experienced the largest decrease in COVID-19 incidence (US Department of Health and Human Services / Center for Disease Control, 2021).

2 Our methods are summarized in Appendix 1.

3 Contrary to the rest of Canada, school boards in Québec (the French *commission scolaires* and the English School Board of Montréal) do not have the autonomy to modify provincial back-to-school guidelines to best fit their local situation.

4 https://santemontreal.qc.ca/fileadmin/fichiers/Campagnes/coronavirus/situation-montreal/rapports-etat-evaluation-montreal/COVID19-Situation-Montreal-Arrondissements-VillesLiees.pdf

5 https://data.ontario.ca/dataset?keywords_en=COVID-19

6 https://www.alberta.ca/covid-19-alberta-data.aspx

7 The public health measure introduced in early October that had the greatest impact on COVID-19 weekly incidence was closing of bars and restaurants, which implied a drop in transmission among 20-29 years old.

